# The Effectiveness of Tirzepatide on weight loss in patients with type 2 Diabetes Mellitus

**DOI:** 10.1101/2025.09.19.25336122

**Authors:** MF Olalere, M Khawaja, O Ndukwe, R Khan, K Kumar, A Sreejith, J Sreedharan, J Muttappallymyalil

## Abstract

Obesity is a significant risk factor for Type 2 Diabetes Mellitus (T2DM), with the United Arab Emirates (UAE) exhibiting a high diabetes prevalence, surpassing the global average. Despite this, there is a notable absence of studies in the UAE and the Gulf region evaluating the efficacy of tirzepatide , a potential weight loss solution. The primary objective is to determine the effectiveness of tirzepatide in inducing weight loss among T2DM patients in the UAE. A retrospective cohort analysis was conducted at Thumbay University Hospital, UAE, assessing the efficacy of Tirzepatide in comparison to Semaglutide in weight loss among adult T2DM patients. The study includes patients initiating treatment between September 2022 and September 2023. Data is collected from medical records, and statistical analysis is performed using SPSS 28 software. 41 patients were included in the study that met our inclusion criteria. There is a significant difference in weight loss outcomes between patients treated with tirzepatide and those treated with semaglutide, with a notably higher percentage of individuals experiencing at least 5% weight loss on tirzepatide. Specifically, 95.1% of those taking tirzepatide experienced weight loss, whereas only 75.6% of those on semaglutide did (P<0.05). Additionally, the mean weight loss difference was positive, at 2.53 kilograms, indicating that, on average, the tirzepatide group had a higher mean weight loss than the semaglutide group. Furthermore, data demonstrated that individuals with a last recorded HbA1c of less than or equal to 7 were more likely to achieve >5% weight loss compared to those with HbA1c greater than 7 (80.0% vs.36.8%) (P<0.01). This statistical evidence supports that patients with better glycemic control lost more weight. The study highlights tirzepatide’s superior effectiveness in weight loss compared to a previously used drug among T2DM patients. By exploring tirzepatide’s efficacy in this diverse population, the research has the potential to inform preventive measures against obesity-related health issues beyond diabetes.

## Introduction

Type 2 Diabetes Mellitus (T2DM) is one of the most common metabolic disorders worldwide. It is recognised as a global health problem with approximately 467 million individuals affected by T2DM^1^. The stark contrast between the global prevalence of diabetes, reported at 9.3%, and the notably higher rate of 16.3% in the United Arab Emirates, underscores a distinct regional challenge in managing and addressing the incidence of diabetes^2^.

T2DM is primarily caused by a combination of two main factors: defective insulin secretion by pancreatic β-cells and the inability of insulin-sensitive tissues to respond to insulin. The intricacies of insulin synthesis, release, and tissue response must be meticulously orchestrated to align with metabolic demands. Any disruptions within these mechanisms can give rise to a metabolic imbalance, ultimately contributing to the development of T2DM^3^.The onset and progression of T2DM bring about a substantial impact on the overall health and well-being of affected patients such as Its association with a reduced life expectancy owing to a greater risk of obesity, heart disease, stroke, peripheral neuropathy and renal disease^4^.

Lifestyle adjustments, such as altering dietary habits and engaging in more physical activity, can prove highly beneficial in enhancing glycemic control with studies showing that exercise can improve glycemic control (lower HbA1C level by 0.66%), with or without significant decrease in body weight. Despite this, the majority of individuals diagnosed with Type 2 Diabetes (T2DM) will necessitate the use of medications for sustained glycemic management over an extended period. Currently, there are many classes of orally available pharmacological agents to treat T2DM: 1) sulfonylureas, 2) meglitinides, 3) metformin (a biguanide), 4) thiazolidinediones (TZDs), 5) alpha glucosidase inhibitors, 6) dipeptidyl peptidase IV (DPP-4) inhibitors, 7) bile acid sequestrants, 8) dopamine agonists, 9) sodium-glucose transport protein 2 (SGLT2) inhibitors and 10) glucagon like peptide 1 (GLP-1) receptor agonists^5^. These diverse classes of antidiabetic medications primarily act by either enhancing insulin secretion, improving insulin sensitivity in peripheral tissues, inhibiting hepatic glucose production, or modulating glucose absorption, collectively aiming to regulate blood glucose levels in individuals with Type 2 Diabetes^6^. Although there are benefits to this, there are challenges and limitations of current conventional medication of T2DM. Sulfonylureas have shown to lose effectiveness (secondary failure), caused by an exacerbation of islet dysfunction with beta cell failure. As a result, the percentage of patients maintaining adequate glycaemic control decreases progressively. In combination with decreased efficacy both sulfonylureas and insulin have shown higher incidence of hypoglycemia. Furthermore, many of these drugs have shown to cause significant weight gain such as: thiazolidinediones, sulfonylureas and meglitinides^7^. The major adverse effects of traditional T2DM medications—most notably obesity—not only increase the risk factors for the disease but also highlight the urgent need for new drugs that successfully address these issues and improve glycemic control and general health.

A major risk factor for developing glucose intolerance is obesity. Those who are obese, particularly those with large amounts of abdominal fat, are prone to a condition called insulin resistance. The pancreas secretes the hormone insulin, which helps cells to absorb glucose and use it as fuel. Insulin helps control blood sugar levels. When cells lack the ability to respond to insulin, they are unable to absorb glucose from the bloodstream as efficiently, which raises blood sugar levels. T2DM results from the pancreas’s inability to produce enough insulin over time due to an increase in demand. Additionally, obesity is often associated with unhealthy lifestyle factors like poor diet and sedentary behavior, which further contribute to the development of diabetes^8^. Prevalence of prediabetes and diabetes has significantly increased in the Gulf countries as a result of the high rate of obesity. In the majority of the GCC countries, the International Diabetes Federation (IDF) found high prevalence rates of obesity in 15% and more than 20% for prediabetic and diabetic patients, respectively^9^. Given that obesity is the key pathophysiologic driver of T2DM, pharmacotherapies that target weight-loss and hyperglycemia may be useful for the management of prediabetes or early T2DM such as trizepatide^10^. Tirzepatide, sold under the brand name Mounjaro, is a single molecule drug, for type 2 diabetic mellitus patient by the combining both glucagon-like peptide-1 (GLP-1) receptor and glucose-dependent insulinotropic polypeptide (GIP)^11^.These are two of the main incretin hormones. When blood sugar levels start to rise after someone eats, this hormone produces more insulin, leading to lowering of blood sugar levels. These dietary stimulated levels of GLP - 1 are unfortunately reduced in T2DM, hence the help of the GLP -1 agonists which could induce the insulinotropic effects. Furthermore, GLP -1 agonists also increase satiety effect^12^. The GIP receptor, found in white adipose tissue, plays a role in regulating lipid metabolism. Activation of the GIP receptor is believed to enhance the ability of adipocytes (fat cells) to efficiently clear dietary triglycerides (TAG) from the bloodstream, while also promoting long-term storage of lipids by supporting the healthy growth of white adipose tissue. Additionally, GIP activity in the central nervous system may provide metabolic benefits by reducing energy expenditure and improving lipid management in peripheral tissues, especially when combined with GLP-1 activity. Conversely, when given together, giving GLP -1 and GIP therapy improves glucose control through the double action on the pancreatic cells leading to the enhancement of insulin secretion^13^.

With the introduction of tirzepatide, it forms the model anti-diabetic medication, showing appealing results in lowering elevated glucose levels, promoting weight loss and lowering the risk of hypoglycemia while offering cardiovascular benefits^14^.This was proven by SURPASS clinical trials which were both multicentric, international, that looked at once weekly doses showed that tirzepatide was found to be superior to other treatment option, showing significant reduction in not only Hba1c levels but in weight loss. Additionally, showed evidence of improvement in cardiovascular risk factors, lipid profiles and reducing non - alcoholic fatty liver disease (NAFLD) biomarkers in the SURPASS - 4 clinical trial^15^. Although we have semaglutide, sold under popular brand name, Ozempic, which is a GLP-1 agonist which has been proven to decrease obesity in patients with T2DM, Fries et al. study revealed that tirzepatide was not only inferior to semaglutide but also far superior in meeting greater weight loss targets and glycemic control^16^.

With the soaring prevalence of diabetes in the UAE, a condition intricately linked to an increased susceptibility to various comorbidities, it is imperative to conduct research on the effectiveness of tirzepatide. This study seeks to explore how tirzepatide, with its potential to aid in weight loss and improve HbA1c levels, could serve as a pivotal intervention in mitigating the associated risks of T2DM. In a country with a prevalence of T2DM double the global percentage and with obesity, a risk factor for T2DM, at 17.8% of the population the need for the effectiveness of tirzepatide in UAE is imperative^17^. This proves the need for evidence - based intervention such as tirzepatide, to help combat obesity in patients of type 2 diabetes to reduce the risk of complications of T2DM.

The purpose of this research is to evaluate how well tirzepatide works in the UAE to help individuals with Type 2 diabetes lose weight. We want to make a significant contribution to the field of diabetes management by analysing its efficacy. Knowing how tirzepatide affects weight loss not only helps patients better manage this disease but also educates policymakers and healthcare professionals about cutting-edge diabetes management strategies. The study’s conclusions have a big impact on how patients are treated since they offer evidence-based methods for enhancing patient outcomes and making healthcare treatments more effective for individuals with Type 2 diabetes. This may open up opportunities for legislators to create regulations that allow users of tirzepatide to lose weight who are not Type 2 diabetics in the UAE. This research has the potential to expand the range of situations in which medical professionals can prescribe tirzepatide by proving its effectiveness in aiding weight reduction in people without T2DM. With the use of such recommendations, medical professionals would be able to prescribe tirzepatide as a workable solution for weight control in a larger population, helping to prevent and treat health issues associated with obesity. This proactive strategy is in line with initiatives to combat the obesity pandemic and improve public health outcomes.

The urgent need to address the rising incidence of obesity and Type 2 diabetes mellitus (T2DM) in the United Arab Emirates (UAE) and throughout the world is the driving force behind this study. Both obesity and type 2 diabetes increase morbidity, mortality, and healthcare expenses dramatically. Obesity is a key risk factor for the development of T2DM. Even with a wide range of treatment options at our disposal, such as medication, bariatric surgery, and lifestyle adjustments, controlling obesity and type 2 diabetes is still a difficult task, especially in communities where these disorders are more prevalent, such as those in the United Arab Emirates.

Although tirzepatide has proven to be an impressive pharmacological option in reduction of not only weight loss but Hba1c , there is a lack of knowledge on its efficacy, particularly with regard to the people of the United Arab Emirates. It is crucial to assess tirzepatide’s effectiveness in this group because of the distinct cultural, culinary, and lifestyle characteristics that are common in the United Arab Emirates.

Additionally, investigating tirzepatide’s possible advantages outside of its well-established application in the treatment of type 2 diabetes may offer insightful information on its wider applicability for weight control. If demonstrated to be successful, tirzepatide may provide a non-invasive, pharmaceutical method of weight loss, which might lessen the burden of comorbidities associated with obesity.

Therefore, by examining the impact of tirzepatide on weight reduction among T2DM patients in the UAE, this study seeks to close the research gap that currently exists. With the ultimate objective of guiding evidence-based programs and recommendations for obesity management in the UAE and abroad, it also aims to investigate the possible effects of tirzepatide for weight control in people without type 2 diabetes.

The primary objective is to determine the effectiveness of tirzepatide on weight loss amongst patients with Type 2 Diabetes Mellitus in the UAE. This study also addresses several secondary objectives such as to determine the proportion of patients who have achieved glycemic control by taking tirzepatide, assessing the prevalence of Albuminuria in patients with T2DM taking tirzepatide and the proportion of patients with Diabetic Retinopathy amongst patients with T2DM taking Tirezepatide. These secondary objectives are designed to explore greater facets of how T2DM can affect many body systems and the importance of medication to control other clinical manifestations related to T2DM.

## Materials and Methods

The research design was a retrospective cohort study. Adult patients with Type 2 Diabetes Mellitus visiting the out-patient department of Internal Medicine during the period from September 2022 to September 2023 will be recruited based on the inclusion criteria. The Study was conducted at Thumbay University Hospital, Ajman, UAE observing the population for 10 months.

The inclusion criteria for this study encompass patients diagnosed with Type 2 Diabetes Mellitus who were prescribed Tirzepatide or Semaglutide and began treatment at the hospital between September 2022 and September 2023. Eligible participants must have been on treatment for a duration of at least 3 months, and both male and female patients of any nationality were considered. Additionally, participants had to be 18 years of age or older. Exclusion criteria include records with incomplete data, as these were not deemed suitable for inclusion in the analysis.

The study instrument used was existing medical records of patients who had been prescribed tirzepatide as part of their diabetes management. Patients prescribed semaglutide were recruited to the study, to compare which drug had the best effect on weight loss. The checklist, approved by experts, was used appropriately to assess our objectives and see if patients met our criteria. A pilot study was conducted involving a small sample of patients who met the inclusion criteria to provide preliminary data for the study. Succeeding the pilot study we then conducted the final study, after which we observed the trends.

The proposal was submitted to the Institutional Review Board (IRB) of Gulf Medical University, and no issues regarding patient confidentiality or safety were raised. Anonymity was ensured by not recording identifying information, and confidentiality will be maintained by storing the document in the Department of Community Medicine for three years, accessible only to researchers, IRB members, and the statistician with appropriate clearance. This research posed no physical, psychological, or social risk to participants and did not involve administering any drugs, food, or placebos.

We used medical records to select patients who met the inclusion criteria, collecting data from September 2022 to September 2023. Approval was obtained from the Institutional Review Board and the Director of Thumbay University Hospital. An Excel sheet was used to record patient demographics (age, gender), medical history (duration of diabetes, comorbidities, prior medications), and details on Tirzepatide and Semaglutide treatment (dosage, duration). The sheet also captured data on concomitant diabetes medications, adjustments made, and patients’ weight and height measurements at regular intervals. Data from patients on both drugs for 3 to 6 months was collected.

The data will be stored for 3 years in the Community Medicine Department as per the GMU policy.The data is recorded as tables and figures by making use of Microsoft Excel and SPSS Statistics Version 27. Additionally, the data is analyzed using the Chi-Square Test for Association as well as a multiple logistic regression to assess the factors. A p-value that is less than 0.05 (P < 0.05) will be statistically significant.

## Results

**Table No. 1** This present study included 41 patients of the total, the majority (65.9%) were males and the remaining were females. The majority of individuals (56.1 percent) in the research are non-Arab, while 43.9 percent are Arab.The majority of individuals (63.4%) fall into the age group of 44 years or younger (<=44).The remaining individuals (36.6%) are in the age group older than 44 (>44). The details are given in table 1.

**Table No. 1.**
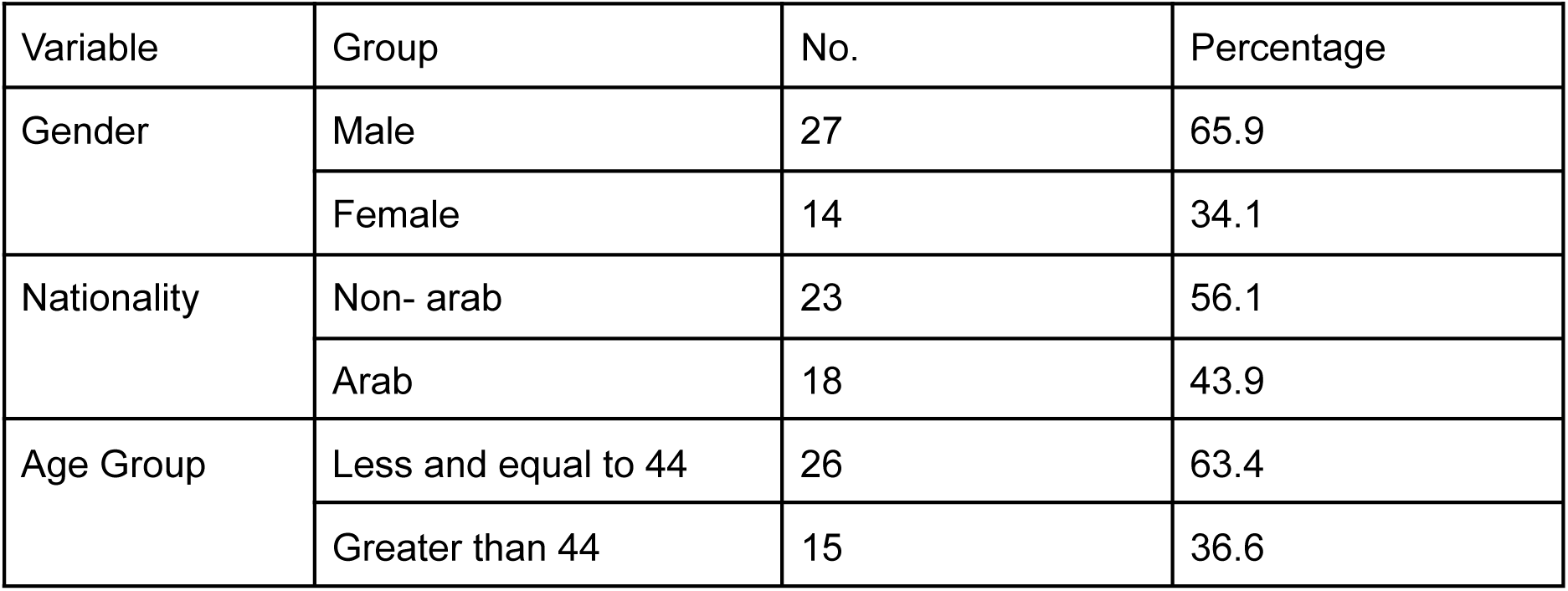
Frequency distribution of gender and ethnicity.

**Table No. 2** The presented data provides insights into various health indicators among the studied group. In terms of retinopathy, 65.9 percent of individuals did not exhibit signs of retinopathy, while 34.1 percent showed its presence. Regarding albuminuria, 70.7 percent of individuals were free from this condition, while 29.3 percent experienced albuminuria. Analysis of the first recorded HbA1c levels revealed that 87.8 percent of individuals had levels exceeding 7, indicating a substantial majority with elevated HbA1c at the outset. Similarly, the last recorded HbA1c levels indicated that 51.2 percent of individuals had levels surpassing 7, underscoring a notable portion with elevated HbA1c towards the end of the study. Lastly, in terms of achieving the target HbA1c, 48.8 percent of individuals successfully reached the target, while 51.2 percent did not.

**Table No. 2.** Frequency distribution of Presence of Retinopathy and Albuminuria, First and Last Recordings of HbA1c and Whether Target Hba1c was Reached in All Patients.

| Variable | Group | No. | % |
| --- | --- | --- | --- |
| Presence of Retinopathy | No | 27 | 65.9 |
|  | Yes | 14 | 34.1 |
| Presence of Albuminuria | No | 29 | 70.7 |
|  | Yes | 12 | 29.3 |
| First recorded HbA1c level | <=7 | 5 | 12.2 |
|  | >7 | 36 | 87.8 |
| Last Recorded Hba1c level | <=7 | 20 | 47.8 |
|  | >7 | 21 | 51.2 |
| Target HbA1c reached | No | 21 | 51.2 |
|  | Yes | 20 | 48.8 |

**Table No. 3** In Dosage 1, 39 percent of individuals were prescribed a dosage of 2.5 mg, while 61 percent received a dosage of 5 mg. For Dosage 2, the majority, at 68.3 percent, received 5 mg, and a smaller proportion (26.8 percent) received 2.5 mg. Dosage 3 shows a diverse prescription pattern, with 68.3 percent receiving 5 mg, 17.1 percent receiving 7.5 mg, and 9.8 percent receiving 10 mg. Dosage 4 indicates a prevalent prescription of 5 mg (46.3 percent), followed by 19.5 percent receiving 7.5 mg, and 7.3 percent receiving 10 mg. Lastly, Dosage 5 reveals a distribution where 34.1 percent were prescribed 5 mg, 14.6 percent were prescribed 7.5 mg, and 7.3 percent were prescribed 10 mg. The table also notes instances of missing data denoted as “Missing 888” across different dosages. Table 3 gives a comprehensive overview of the dosages administered within each visit from the patient.

**Table No. 3.** Tirzepatide Dosage Distribution Across Each Visit.

| Variable | Dosage (mg) | No. | % |
| --- | --- | --- | --- |
| Dosage 1 | 2.5 | 16 | 39 |
|  | 5 | 25 | 61 |
| Dosage 2 | 2.5 | 11 | 26.8 |
|  | 5 | 28 | 68.3 |
|  | 7.5 | 2 | 4.9 |
| Dosage 3 | 2.5 | 2 | 4.9 |
|  | 5 | 28 | 68.3 |
|  | 7.5 | 7 | 17.1 |
|  | 10 | 4 | 9.8 |
| Dosage 4 | 2.5 | 1 | 2.4 |
|  | 5 | 20 | 46.3 |
|  | 7.5 | 8 | 19.5 |
|  | 10 | 3 | 7.3 |
|  | Missing 888 | 9 | 22 |
| Dosage 5 | 2.5 | 1 | 2.4 |
|  | 5 | 14 | 34.1 |
|  | 7.5 | 7.3 | 14.6 |
|  | 10 | 7.3 | 7.3 |
|  | Missing 888 | 17 | 41.5 |

**Table No. 4.** On the first visit, the majority of patients were placed on medication for 56 days (68.3%). The 2nd visit shows that 31.7 percent of individuals had a treatment duration of 28 days, 51.2 percent had a duration of 56 days, and 17.1 percent had a duration of 84 days. The 3rd visit, 29.3 percent of individuals had a treatment duration of 28 days, 46.3 percent had a duration of 56 days, 2.4 percent had a duration of 58 days , and 22.0 percent had a duration of 84 days. The 4th visit shows that 46 percent had a treatment duration of 28 days, 51.2 percent had a duration of 56 days, and 12.2 percent had a duration of 84 days. The 5th visit indicates that the majority of patients were prescribed the to be on the medication for 56 days (46.3%).

**Table No. 4.** Tirzepatide Prescription Duration.

| Variable | Days | No. | % |
| --- | --- | --- | --- |
| Duration of Treatment 1 | 28 | 11 | 26.8 |
|  | 56 | 28 | 68.3 |
|  | 84 | 2 | 4.9 |
| Duration of Treatment 2 | 28 | 13 | 31.7 |
|  | 56 | 21 | 51.2 |
|  | 84 | 7 | 17.1 |
| Duration of Treatment 3 | 28 | 12 | 29.3 |
|  | 56 | 20 | 49 |
|  | 84 | 9 | 22 |
| Duration of Treatment 4 | 28 | 6 | 14.6 |
|  | 56 | 21 | 51.2 |
|  | 84 | 5 | 12.2 |
|  | Missing | 9 | 22 |
| Duration of Treatment 5 | 28 | 3 | 7.3 |
|  | 56 | 19 | 46.3 |
|  | 84 | 2 | 4.9 |
|  | Missing | 17 | 41.5 |

**Table No. 5** The data below shows that amongst individuals on Tirzepatide whose last Hba1C recorded was less than or equal to 7, 20 percent achieved a weight loss of less than/equal to 5 percent and 80 percent achieved a weight loss of greater than 5 percent. Individuals whose last Hba1C recorded was less than or equal to 7 are more likely to achieve >5% weight loss compared to those with HbA1c greater than 7 (80.0% vs. 36.8%). There is statistical evidence to support the association between the level of HbA1C that was recorded last and the percentage of weight loss (p = 0.005).

**Table No. 5.**
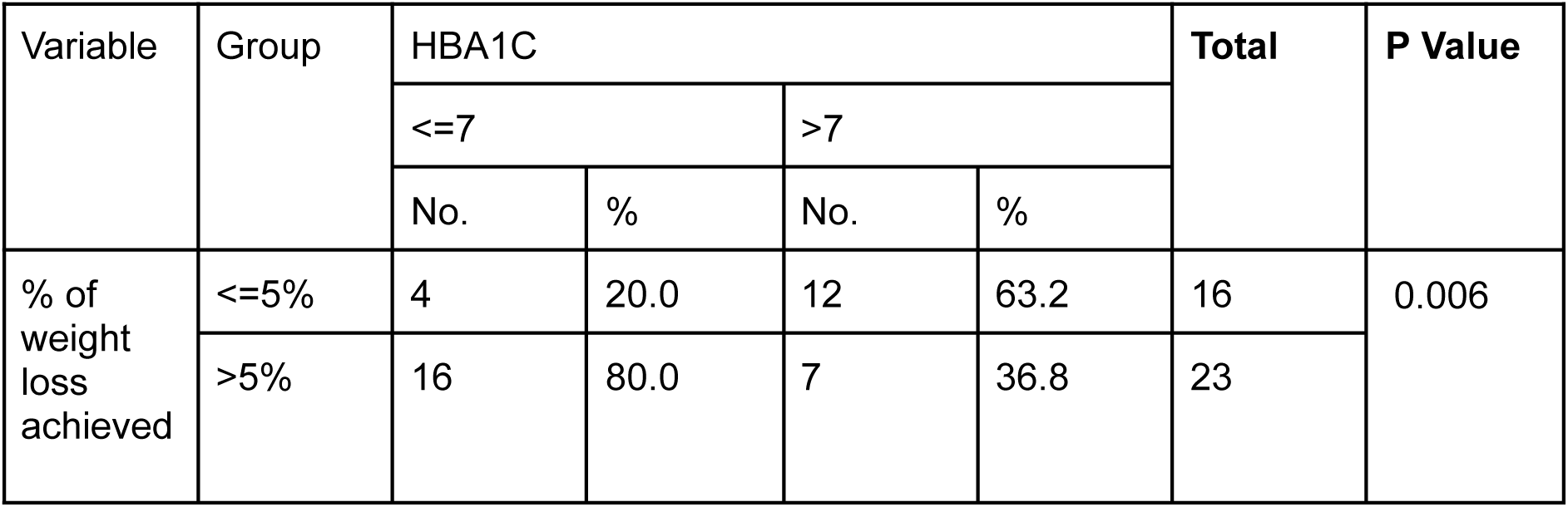
Association Between HbA1C Levels and Weight Loss Achievement.

| Variable | Group | HBA1C |  |  |  | Total | P Value |
| --- | --- | --- | --- | --- | --- | --- | --- |
|  |  | <=7 |  | >7 |  |  |  |
|  |  | No. | % | No. | % |  |  |
| % of weight loss achieved | <=5% | 4 | 20.0 | 12 | 63.2 | 16 | 0.006 |
|  | >5% | 16 | 80.0 | 7 | 36.8 | 23 |  |

**Table No. 6** The crosstabulation table shows the distribution of individuals across the different levels of tirzepatide and semaglutide with their respective outcomes on whether individuals lost weight or not. The crosstabulation table reveals a notable association between the type of drug administered (Tirzepatide or Semaglutide) and the occurrence of weight loss. Among individuals taking Tirzepatide, 95.1% experienced weight loss, while 75.6% of those on Semaglutide.

**Table No. 6.** Association Between Drug and Weight Loss Achievement.

| Variable | Group | Weight loss (yes/no) |  |  |  | Total | P Value |
| --- | --- | --- | --- | --- | --- | --- | --- |
|  |  | No |  | Yes |  |  |  |
|  |  | No. | % | No. | % |  |  |
| Drug | Tirzepatide | 2 | 4.9 | 39 | 95.1 | 41 | 0.012 |
|  | Semaglutide | 10 | 24.4 | 31 | 75.6 | 41 |  |

**Table No. 7** The positive mean difference in the amount of weight loss indicates that, on average, the group associated with Tirzepatide had a higher mean weight loss compared to the group associated with Semaglutide. The confidence interval further supports this by providing a range of values within which we are 95% confident that the true mean difference lies, and it is entirely positive.

**Table No. 7.** Comparison of Weight Loss Means with t-Test Analysis.

|  |  | t-Test for Equality of Means |  |  |  | 95% Confidence Interval of Difference |  |
| --- | --- | --- | --- | --- | --- | --- | --- |
|  |  | Significance One - Sided p | Significance Two sided p | Mean Difference | Std Error Difference | Lower | Upper |
| **Amount of weight loss** | Equal variances assumed | 0.012 | 0.023 | 2.53 | 1.09 | 0.344 | 4.71 |
|  | Equal variances not assumed | 0.010 | 0.020 | 2.53 | 1.06 | 0.411 | 4.65 |

**Table No. 8** The table shows the effect size which helps to understand how big of a difference there is between the two groups (tirzepatide and semaglutide) in terms of weight loss.All three effect size measures (Cohen’s d, Hedges’ correction, and Glass’s delta) consistently indicate a large effect size, emphasizing the substantial difference in weight loss amounts between the tirzepatide and semaglutide groups. All three measures agree that the difference in weight loss between tirzepatide and semaglutide is not only statistically significant (meaning it’s not just due to chance) but also practically significant, indicating a substantial difference in the amount of weight lost.

**Table No. 8.** Effect Size Measures for Weight Loss.

|  |  |  |  | 95% Confidence Interval of Difference |  |
| --- | --- | --- | --- | --- | --- |
|  |  | Standardizer | Point Estimate | Lower | Upper |
| **Amount of weight loss** | Cohen's d | 4.53 | 0.448 | 0.076 | 1.037 |
|  | Hedges' correction | 4.58 | 0.552 | 0.075 | 1.026 |
|  | Glass's delta | 3.94 | 0.643 | 0.140 | 1.137 |

## Discussion

This study provides insight and information into how successful tirzepatide is as a powerful weight loss option for patients with Type 2 Diabetes Mellitus. With high rates of obesity and T2DM within the UAE, the region provides an excellent background to showcase how efficient and practical the use of tirzepatide is for weight loss. In order to prevent and manage T2DM, treating obesity is essential. Reducing body weight helps prevent, manage, and cause remission of diabetes in some instances. Lifestyle changes and other pharmacological options have been the mainstay treatment for those who are overweight and obese with T2DM, however the emergence of tirzepatide has proven to be better than its counterparts, hence why this study chooses to examine its effectiveness in the UAE.

Tirzepatide, dual GLP-1 and GIP agonist, in this study showed to reduce body weight significantly, compared to semaglutide, a GLP agonist. Body weight reduction was defined as more than 5% of the initial weight of the patient. 95.1% of those taking Tirzepatide experienced weight loss, whereas only 75.6% of those on Semaglutide did (P<0.05). These results are similar to other studies which showed that using tirzepatide resulted in a weight loss of 17.8% (95% CI: 16.3%-19.3%) compared with 12.4% (95% CI: 11.5%-13.4%) for semaglutide^41^. Furthermore, while our study yielded favorable results, it’s noteworthy to mention that the dosages of both tirzepatide and semaglutide differed from our own protocol. While the semaglutide dosage remained the same, patients on tirzepatide underwent a gradual dosage escalation. This is the standard guideline recommended by the Food and Drug Administration (FDA) and European Medicine Agency (EMA) commencing with 2.5 mg once weekly with subsequent adjustments tailored to individual responses after four weeks, with maximum dose at 15mg ^42^.This is also corroborated by the SURPASS trials which also utilised this escalation algorithm for tirzepatide, while obtaining increased reduction in weight loss compared to placebo and semaglutide^43^. Furthermore, the average weight loss difference was positive, measuring 2.53 kg, meaning that the tirzepatide group lost more weight on average than the semaglutide group. This echoes findings from a similar study, demonstrating a significant mean difference of 6.33 kg, between treatment and control group suggesting that tirzepatide exhibits greater efficacy in promoting weight loss compared to semaglutide^44^. Notably, the patients with lower Hba1c achieved better weight loss reduction with tirzepatide, highlighting the drug’s dual efficacy in addressing both weight management and glycemic control in patients. The research showed that data demonstrated that individuals with a last recorded HbA1c of less than or equal to 7 were more likely to achieve >5% weight loss compared to those with HbA1c greater than 7 (80.0% vs. 36.8%) (P<0.01). This statistical evidence supports that patients with better glycemic control lost more weight.

Though tirzepatide and semaglutide have the potential to reduce weight, minor variations in how they interact with the body’s metabolic and physiological pathways can affect both their overall efficacy and specific to patients tolerability. Semaglitude does have more research behind it, with it gaining FDA approval in 2017, while tirzepatide in 2022^45^.When tirzepatide and semaglutide were compared for weight loss, tirzepatide had more notable weight loss outcomes. This is probably because, in contrast to semaglutide, which exclusively targets the glucagon-like peptide 1 (GLP1) receptor, tirzepatide targets both the glucose-dependent insulinotropic polypeptide receptor and GLP-1.

Additionally data showed that amongst 41 patients, 20 patients (48.9%) met the Hba1c target of 7. Conversely, a study showed that more trizepatide-treated patients achieved an HbA1C of <7% and a more robust reduction in fasting serum glucose^46^. 29.3% and 34.1% had albuminuria and retinopathy, respectively. Sana et al. reported 31.56% of diabetic patients with microalbuminuria, indicating a higher occurrence compared to our own research. Notably, the presence of albumin in urine serves as an indicator of glomerular involvement in type 2 diabetes mellitus (T2DM), signalling the onset of diabetic nephropathy. It underscores the importance of stringent glycemic control as a preventive measure to delay the onset of microalbuminuria and other diabetic complications^47^. Not only this but tirzepatide has shown a beneficial effect on albuminuria level in systematic reviews^48^. With diabetic retinopathy being one of the major chronic microvascular complications of T2DM and 35.0% of diabetics have a degree of retinopathy, hence why diabetic eye screening is essential.

Initial screening is crucial to detect any pathological changes, followed by annual check-ups to monitor the progression of diabetic retinopathy. Although diabetic retinopathy may not be fully reversible, diligent diabetes care with intensive blood glucose control can effectively delay its progression^49^.

Within this study it is imperative to acknowledge both the strengths and limitations inherent to its design and methodology. One strength is the study design, a retrospective cohort , we were able to make use of pre-existing data from medical records which saved not only time but resources that would otherwise be needed for data gathering.Despite these strengths, there were some challenges with this research. One of them is that the sample size remained relatively small, which may have limited our ability to detect enough association within our data. However, we hypothesize this is due to patients not meeting the inclusion criteria of being on tirzepatide for a sufficient amount of time for the trial. This may be due to the limited availability of the drug at the pharmacy, insurance and payment issues as well as patients having difficulty administering the drug. The last one is that the incremental dosage adjustments, increasing by 2.5 units per visit, may have contributed to a limitation in our ability to discern further efficacy of the drug, although this nature in prescribing is not used in many other trials and research on tirzepatide but it is advised by the FDA.

Medication for glycemic management and weight loss, such as tirzepatide and semaglutide, is beneficial, but works better when combined with a healthy diet and frequent exercise. The foundation of diabetes care is exercise, which promotes improvements in glucose utilization, insulin sensitivity, and weight control. Including regular exercise in everyday activities lowers cardiovascular risk factors, improves glycemic control, and enhances general health and well-being^50^. Similarly, maintaining optimal metabolic health and managing diabetes mellitus requires sticking to a healthy diet full of fruits, vegetables, whole grains, and lean meats. A balanced diet lowers the risk of complications from diabetes, encourages weight loss or maintenance, and helps control blood glucose levels. Tirezepatide was approved by the FDA as an adjunct to exercise and dietary adjustments so that patients with T2DM can experience synergistic advantages that enhance their overall health outcomes, including glycemic control and weight management^51^.

Being overweight or obese raises not only your risk of developing T2DM, but also other conditions such as: arterial hypertension, coronary heart disease and cerebral vasculopathy. Obesity is a serious public health issue associated with increased morbidity and mortality^52^. When managing obesity, it is important to take a multifaceted strategy such as using medication, behavioural therapies, exercise management, food adjustments, and, if necessary, surgery^53^. Pharmacologically, FDA has only approved five medications—orlistat, phentermine/topiramate, naltrexone/bupropion, liraglutide (3 mg), and semaglutide 2.4 mg—as substitutes for the treatment of obesity^54^. With semaglutide 2.4 mg, exhibiting greater efficacy compared with previously existing pharmacological options^55^. Research has been conducted on tirzepatide being used as a treatment for obesity, with a recent study showing tirzepatide to have a greater reduction in body weight compared to the control group (semaglutide) [MD = −5.65, 95% CI (−7.47, −3.82), p < 0.001]. Additionally, it has served to reduce waist circumference and BMI^56^. Encouragingly, in November 2023, FDA has approved Zepbound, for chronic weight management in adults diagnosed with obesity, overweight, or weight-related conditions (T2DM or high blood pressure). Tirzepatide, which is the active ingredient in Zepbound, is already approved as Mounjaro for glycemic control in T2DM. Zepbound in studies has shown that greater proportions of patients who received Zepbound achieved at least 5% weight reduction compared to placebo^57^. Although Mounjaro has only been approved in the UAE for better glycemic control for weight loss, studies have not only shown its benefits in effectively losing weight but reveals presents a promising avenue for weight loss in individuals with obesity, potentially serving as a proactive measure to mitigate the development of associated comorbidities, which would be advantageous in population with high obesity prevalence such as the UAE.

## Conclusion

Our study investigated the effectiveness of tirzepatide ( 2.5, 5, 10, 15mg) compared to semaglutide (1mg) in weight loss in patients with T2DM.Through rigorous analysis, tirzepatide demonstrated superior effectiveness in promoting weight loss, surpassing the outcomes observed with semaglutide. Achieving greater weight reduction, shows a compelling advantage in the treatment of diabetes mellitus. Weight loss plays a pivotal role in improving glycemic control and metabolic parameters in individuals with diabetes mellitus, leading to reduced insulin resistance, improved beta-cell function, and decreased cardiovascular risk factors.

Moreover, our study has important implications for clinical practice and patient care. The effectiveness of tirzepatide in weight loss and reduction in Hba1c underscores its potential as a promising therapeutic option for individuals with diabetes mellitus, particularly those within the population of the UAE, who are more likely to be diagnosed with this chronic disease.

However, it is important to acknowledge the limitations of our study, including small sample size. Further research is warranted to show which particular dose of tirzepatide is more effective in weight loss and in which dosage shows more promise in greater weight loss in those who are either overweight or obese. This study can be used to by policy makers and regulatory borders to explore the greater use of tirzepatide for obesity and obesity related conditions, Overall, our findings contribute to the growing body of evidence on tirzepatide and its role in T2DM management. By advancing our understanding of its efficacy and use, we can better inform clinical decision-making and optimize patient outcomes in the treatment of diabetes mellitus.

## Data Availability

All data produced in the present study are available from the corresponding author upon reasonable request.

## Notes

### Competing Interest Statement

The authors have declared no competing interest.

### Funding Statement

This study did not receive any specific funding.

### Author Declarations

Ethics committee/IRB of Gulf Medical University gave ethical approval for this work. All patient data were anonymized prior to analysis, and no identifiable information was included.

